# A systematic review of the methodological considerations in *Campylobacter* burden of disease studies

**DOI:** 10.1101/2024.11.08.24316954

**Authors:** Megan Tumulty, Carlotta Di Bari, Brecht Devleesschauwer, Sara M. Pires, Zubair Kabir

## Abstract

**Background:** Campylobacteriosis is a major zoonotic and foodborne disease (FBD), posing a substantial social and health economic burden on human health. Burden of disease (BoD) studies, which increasingly use the disability-adjusted life years (DALYs) metric, provide comprehensive insights into disease effects. However, the complexity of DALY calculations, combined with diverse causative agents and research gaps, complicates cross-regional comparisons. This review evaluates existing *Campylobacter* BoD studies and interrogates their methodological approaches and assumptions in quantifying DALYs.

**Methods/Principal Findings:** A systematic search of PubMed, EMBASE, Web of Science, and selected grey literature databases was conducted to identify existing *Campylobacter* BoD studies. Studies assessing the BoD methodology and calculation using the DALY framework were considered. In total, 23 studies met the predefined inclusion criteria. Of these, 19 were single-country studies, while 4 were multi-country analyses. A significant data gap exists, with limited or no studies from low- and middle-income countries, exemplified by just one study obtained from Rwanda. Most studies used an incidence- and pathogen-based approach to estimate DALYs, excluding social weighting, in line with the Global Burden of Disease (GBD) study. Methodological discrepancies were noted, especially in disability weight (DW) assignment, health state classification, and life expectancy table usage. Most single-country studies (n=8) used national life tables rather than universal ones, challenging cross-country comparisons due to a lack of standardisation.

**Conclusion:** Significant variations in the methodological approaches and assumptions for *Campylobacter* BoD studies exist. Addressing these disparities is essential for harmonising methodological design choices using the DALYs metric to inform evidence-based public health policies and interventions.

**PROSPERO Registration Number:** The protocol for this study was registered with the International Prospective Register of Systematic Reviews (PROSPERO), which can be accessed under the registration number <u>CRD42023414973</u>.

## Introduction

Campylobacteriosis is a significant zoonotic and FBD (1,2), primarily transmitted through contaminated poultry, accounting for up to 80% of human cases (3–6). While most infections are self-limited, relapse is common, and post-infectious sequelae like Guillain-Barré syndrome (GBS) and reactive arthritis (ReA) can occur (7,8). Additionally, it has been hypothesized that *Campylobacter*-induced disease may be linked to functional gastrointestinal disorders, including irritable bowel syndrome (IBS), and inflammatory bowel disease (IBD) (2,8).

The epidemiology of *Campylobacter* infection differs markedly between high-income and low- and middle-income countries (LMICs) (2,9). In high-income nations, human campylobacteriosis is primarily linked to contaminated food (10), with notable incidence rates observed in North America (11), Australia (12), and Europe (10,13). In 2021, it was a leading cause of FBD in Europe, with 127,840 confirmed cases, peaking in late spring and summer (10,14). Globally, campylobacteriosis incidence has increased over the past decade, accounting for over 62% of all reported human zoonotic cases in 2021 (absolute cases increased by 7,296 from the year prior) (2,10). Campylobacteriosis significantly contributes to the global BoD, as evidenced by DALY estimates. The WHO Foodborne Disease Burden Epidemiology Reference Group (WHO/FERG), reported that *Campylobacter* caused 95.6 million foodborne illnesses in 2010, resulting in 2.1 million foodborne DALYs worldwide (15). Since 2007, it has been the most frequently reported FBD in the European Union/European Economic Area (EU/EEA), with approximately 129,960 cases in 2021 (13). However, the discrepancy between reported and projected cases suggests the actual number may approach 9 million annually (16,17), highlighting an underestimation due to incomplete reporting and imprecise estimates (18,19). Conversely, in LMICs, *Campylobacter* infections are often endemic (20–23) and linked to environmental exposures, with disparities in surveillance systems, underreporting, and diagnostics hindering data collection (1,9,18,24).

Understanding the burden of campylobacteriosis is essential for informing effective public health and food safety strategies (25,26). Quantifying DALYs associated with *Campylobacte*r infections enables policymakers to assess health and economic impacts, thereby guiding resource allocation and mitigation efforts (25). DALYs, increasingly utilised in BoD studies, combine premature mortality (Years of Life Lost, YLL) and disability (Years Lived with Disability, YLD) into a single metric (27–29). Introduced in the early 1990s in the GBD study, DALY estimation has been widely adopted in various research, such as the Burden of Communicable Diseases in Europe (BCoDE) and the WHO/FERG (30,31).

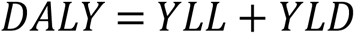

Calculating DALYs involves several methodological choices and assumptions (28,32). However, these methodological variations, along with a lack of transparency, create uncertainties for researchers, policymakers, and institutions, complicating the comparison of BoD estimates across regions (33).

This paper solely focused on the human health burden of campylobacteriosis. The aim of this review was to contribute to the literature by systematically locating, appraising, and summarising the best available evidence from existing *Campylobacter* BoD studies to date, and to comprehensively review the methodological choices undertaken.

## Method

### Search Strategy

This systematic review was carried out in accordance with the Preferred Reporting Items for Systematic Reviews and Meta-Analyses (PRISMA) Statement (S1 PRISMA Checklist) (34). According to the PRISMA guidelines, a flow diagram was used throughout this study to illustrate the step-by-step study selection process, and rationale was provided for those studies that were excluded (34).

Three different search strategies were employed-database search, grey literature, and snowballing. Namely, PubMed, Web of Science, and Embase were systematically searched for potentially eligible studies during week 25, 2023. All studies that quantified DALYs were included, regardless of whether their primary focus was on assessing the BoD or other aspects (e.g., cost-effectiveness and cost of illness studies). The search included the use of the following terms: *Campylobacter,* Campylobacteraceae, burden of disease, disability-adjusted life year, years of life lost, years lived with disability, cost effectiveness and cost of illness. The search strategy employed Boolean operators and Medical Subject Headings (S2 Search Strategy). Additionally, a grey literature search was conducted using Google Scholar and OAIster. Moreover, the search was supplemented using the snowballing search strategy, involving the manual screening of reference lists from included publications to identify additional eligible studies missed during the initial search.

### Eligibility Criteria

Peer-reviewed journal articles and grey literature published between 1990 and 2023 were considered for inclusion. Studies published prior to 1990 were excluded due to the introduction of the DALY metric in the early 1990s (29). All studies utilizing the BoD methodology to calculate DALYs, regardless of their primary focus, were included. Only studies assessing *Campylobacter* BoD in human populations were eligible. There were no language or study type restrictions; however, abstracts, conference proceedings, reviews, letters to editors, and studies without full-text availability were excluded from the search.

### Study Selection and Screening

Titles and abstracts of publications retrieved from preselected databases (PubMed, Web of Science, and Embase), as well as grey literature sources, were organised and managed using the Zotero reference manager, with duplicates identified and removed. The publications were then imported into Rayyan.ai (35) to further manage duplicates and ensure accuracy. Duplicates were identified based on author(s), publication year, and title, with abstracts compared if needed. Subsequently, articles underwent eligibility screening, including evaluation based on title, abstract, and full text, conducted by an independent reviewer (MT), with CDB reviewing. A sample of articles was pilot screened by the study supervisors (CDB and ZK) to assess conformity. Any uncertainties or discrepancies were discussed (MT, CDB, ZK), and documentation of reasons for article rejection provided in a PRISMA flow diagram (Fig 1).

**Fig 1.**
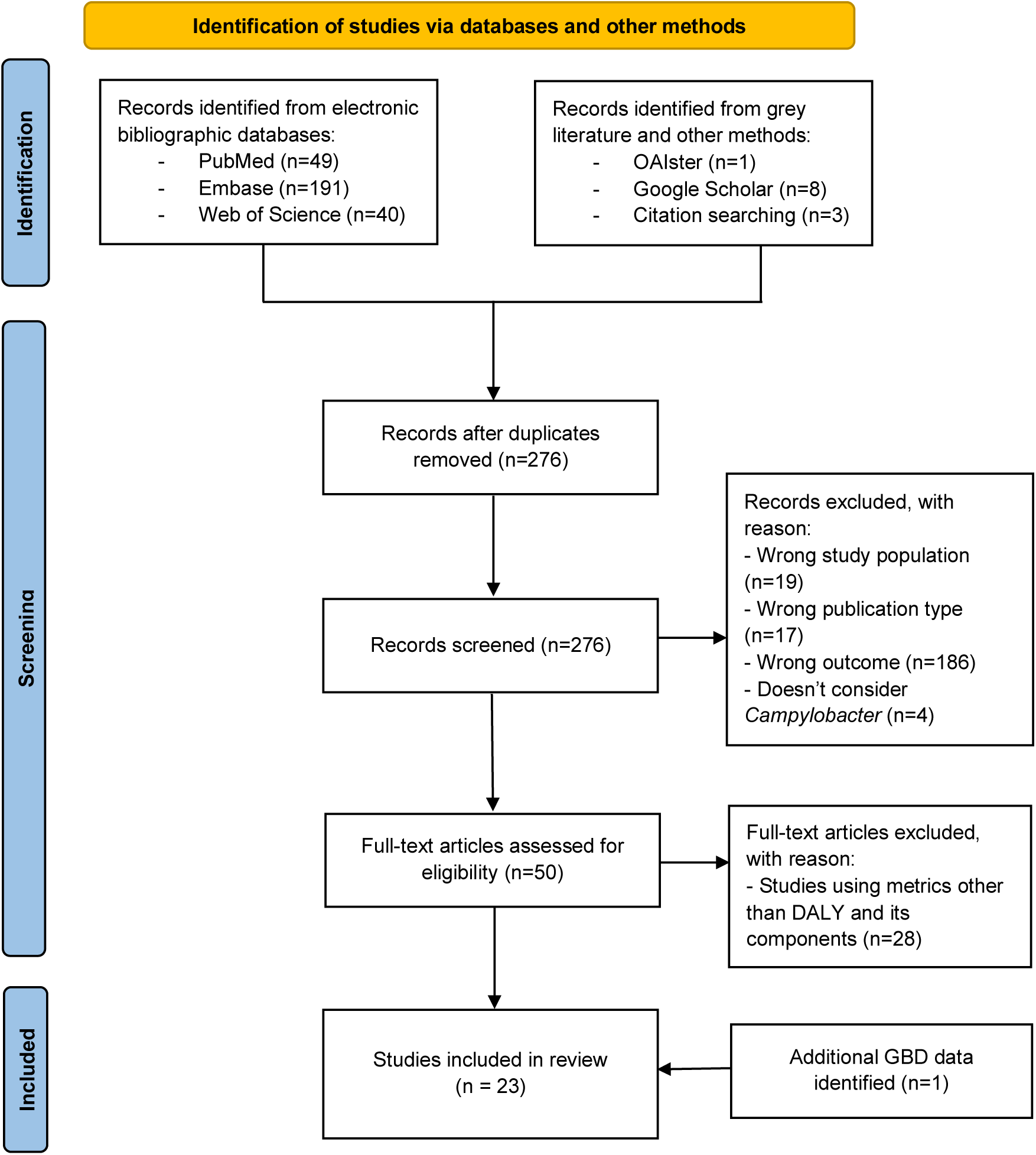
Flow diagram illustrating the literature search and study selection, which has been adapted from the PRISMA 2020 guidelines for systematic reviews (39).

### Data Abstraction

Data abstraction for all eligible studies was conducted by an independent reviewer (MT) through a full text assessment. This process utilized a standardized data abstraction form (S3 Data Abstraction Form), which was piloted with study supervisors and adapted from systematic literature reviews by Charalampous *et al.,* (36) and Di Bari *et al.,* (28) to align with the objectives of this review. The data abstraction form facilitated the collection of study characteristics, including data sources for epidemiological data, DALYs results, and methodological considerations for calculating YLDs and YLLs. YLDs in DALY calculations can be derived from either an incidence- or prevalence-based approach. The incidence perspective, used by BCoDE and WHO/FERG, projects future BoD based on current exposures and focuses on pathogen-related outcomes, including sequelae (28,37,38). On the other hand, the prevalence-based approach, employed by GBD, captures the population’s health status at a specific point in time (25,28,37). GBD also adopts an outcome-based approach to allocate BoD across health conditions and estimate major infectious disease-related outcomes (38). Following data abstraction, a second reviewer (CDB) reviewed the finalized form. Any revisions or discrepancies were resolved through discussion and consensus.

At present, there is a lack of established tools specifically designed to evaluate the quality of studies estimating DALYs. Consequently, as the primary objective of this study is to evaluate the methodological approaches and assumptions employed in *Campylobacter* BoD studies, a comprehensive assessment of overall publication quality was conducted as part of this systematic review. This renders the use of specific quality appraisal tools unnecessary. Furthermore, a meta-analysis was not conducted in this review. This decision was primarily driven by the study’s focus on exploring various methodological choices in *Campylobacter* BoD studies, rather than estimating pooled DALY rates.

## Results

### Literature Search

Fig 1 illustrates a flow diagram detailing the selection process for studies in this systematic review. A total of 292 publications were initially identified through specified databases, grey literature sources, and citation searches. Following the removal of duplicates (n=16), 50 full-text articles were assessed for eligibility. Subsequently, 28 articles were excluded as they utilized health metrics other than YLD, YLL, or DALYs in the BoD assessment. Furthermore, an additional study focusing on GBD data was identified. Finally, data were extracted from 23 BoD studies for analysis.

### Study Characteristics

Of the 23 eligible BoD studies included for review, 19 were performed at a single-country scale, while 4 were multi-country analyses, as depicted in Fig 2. Within the multi-country studies, Mangen *et al.,* (40) and Cassini *et al.,* (31) undertook regional assessments across countries within the EU/EEA. Additionally, 2 studies addressed the burden of campylobacteriosis on a global level, with one conducted by Kirk *et al.,* (15) under the WHO/FERG study. They utilized a pathogen-based approach to calculate DALYs, considering estimated cases, sequelae, and fatalities. Conversely, Vos *et al.,* (41) summarised GBD methodologies, presenting results from an outcome-perspective. Although GBD provides specific estimates for campylobacteriosis, these are often categorised under broader infectious disease groups rather than as a discrete entity. For instance, *Campylobacter* is integrated within overall estimates for diarrhoeal diseases. The study by Vos *et. al.,* (41) did not mention campylobacteriosis independently but included it within the collective analysis of diarrhoeal diseases. The appendix of the article details that during data input, diarrhoeal aetiologies, including *Campylobacter,* were extracted from scientific literature reporting the proportion of diarrhoea cases testing positive for each pathogen (41). This methodology enables comprehensive estimates of diarrhoeal disease burden but obscures discrete data on specific pathogens like *Campylobacter*.

**Fig 2.**
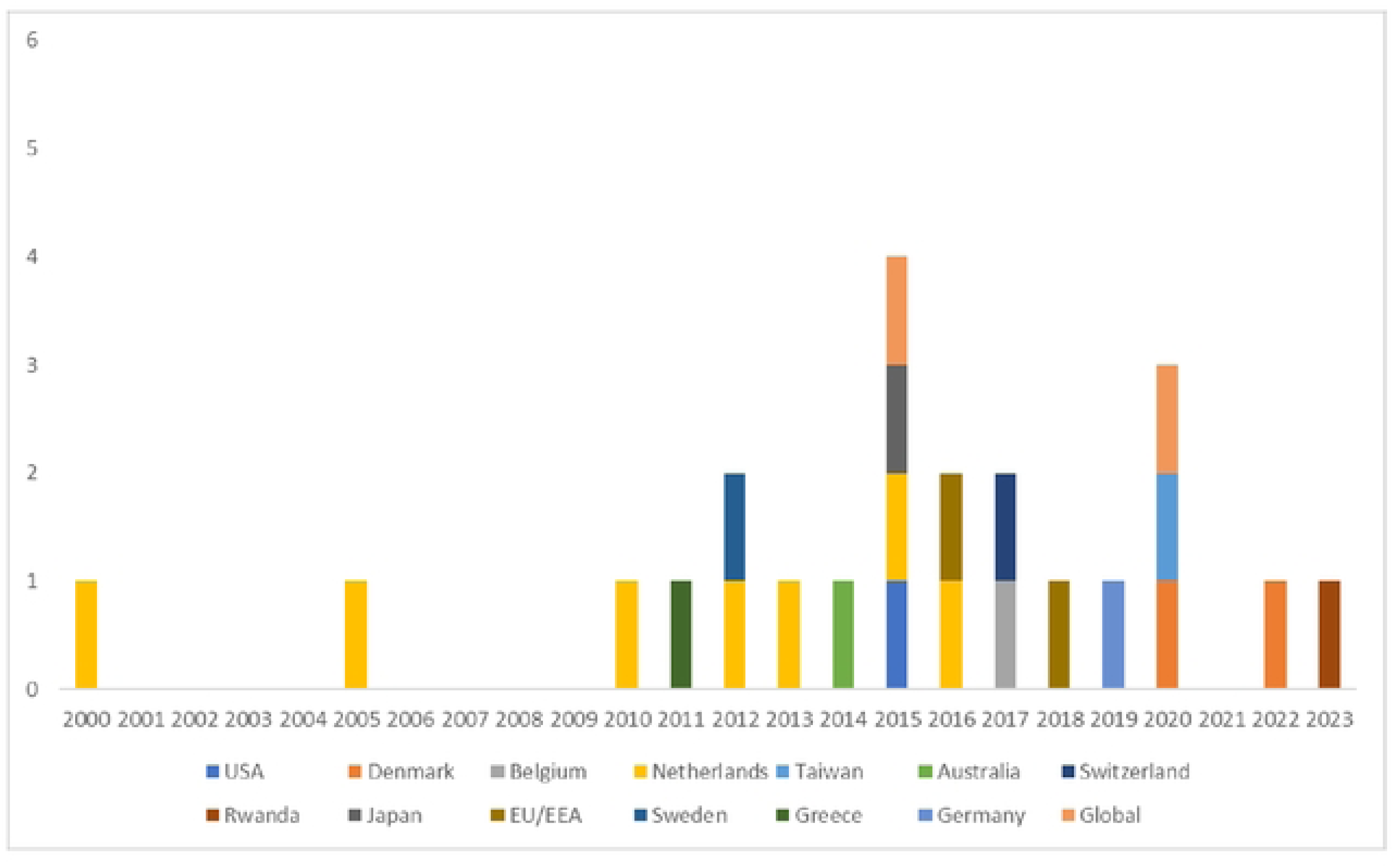
Existing *Campylobacter* BoD studies compiled from published literature based on year of publication.

Over the 2000-2023 period, the largest number of national-level studies originated from the Netherlands (n=7) (42–48), followed by Denmark (n=2) (49,50). Additional individual studies spanned various countries, including the United States (26), Belgium (51), Taiwan (52), Australia (53), Switzerland (54), Japan (55), Sweden (56), Greece (57), German (58), with the most recent publication stemming from Rwanda in 2023 (59).

### Methodological Choices

#### Approaches for Estimating YLL: Choice of Life Expectancy Table

Most studies (n=21) included YLL due to campylobacteriosis. However, one study by Haagsma *et al.,* (45) explicitly omitted the YLL component in the DALY calculation, as they did not associate IBS with increased mortality. Among the single-country independent BoD studies (n=8), national life tables were predominantly utilized to compute YLLs (26,47,50,52–54,56,58). Moreover, four of the independent single-country studies employed the aspirational global Coale-Demeny (West level 25 or 26) model life tables (42–44,48).

Notably, GBD data for campylobacteriosis does not exist as a standalone disease; rather, it is classified as an etiology of overall infectious diseases. Although Vos *et al.,* (41) utilized GBD life tables in their analysis, it is important to clarify that these tables do not provide specific YLL data for campylobacteriosis. The remaining single- or multi-country studies employed various life expectancy tables, including aspirational model life tables akin to those used in the GBD study (31,46,55), standard life model tables from WHO (15,49,59), or a combination of the aforementioned tables (40,51,57).

#### Social Weighting: Age Weighting and Time Discounting

In total, five studies applied either age weighting or time discounting rates, but none of the studies implemented both social weighting parameters (Table 1). Among the studies employing these variables, time discounting was more prevalent, as evidenced by four studies (40,42,44,47). Notably, the discounting rates varied significantly across studies. For instance, Mangen *et al.,* (42) and Havelaar *et al.,* (44) presented disease burden data both undiscounted and discounted at a rate of 1.5%. In contrast, an earlier study by Havelaar *et al.,* (47) in 2005 presented disease burden data using different discounting rates, including undiscounted data and data discounted at a rate of 4%. Similarly, the regional study by Mangen *et al.,* (40), which estimated the burden of campylobacteriosis across six EU/EEA countries, presented undiscounted data alongside different discounting rates, including 3%, 6%, and country-specific rates. Although Gibney *et al.,* (53) did not utilize time discounting, they did apply age weighting rates for acute gastroenteritis to the reference population.

**Table 1.** Summarises these methodological differences, as extracted from the studies included in this review (for the full data abstraction form, refer to S3 Data Abstraction Form).

| Region | Study | General |  | YLL | YLD |  |  |  | Results |
| --- | --- | --- | --- | --- | --- | --- | --- | --- | --- |
|  |  | Time discount | Age weighting | Life table | Approach | Duration source | DW source | Health states | DALYs per case |
| Global | Kirk *et al.*, 2015 (15) | No | No | WHO 2050 | Incidence | Literature | GBD 2010 | Diarrhoea, GBS, Death | 0.022 |
|  | Vos *et al.*, 2020 (41)<sup>a</sup> | - | - | GBD | Prevalence | - | GBD 2019 | - | - |
| Regional | Cassini *et al.*, 2018 (31) | No | No | GBD 2010 | Incidence | Literature | European DW | GE, GBS, ReA, IBS, Death | 0.010 |
|  | Mangen *et al.*, 2016 (40) | Yes | No | West Level 25 and 26; country-specific; and | Incidence | Literature | GBD DW (Mangen *et al.*, 2013) | GE, GBS, ReA, IBS, Death | 0.027 |
|  |  |  |  | GBD 2010 |  |  |  |  |  |
| National | Scallan *et al.*, 2015 (26) | Unknown | Unknown | Country-specific | Incidence | Unknown | Dutch DW | GE, GBS, ReA, IBD, IBS, Death | - |
|  | Pires *et al.*, 2020 (49) | No | No | WHO life tables | Incidence | Data | GBD 2013 | Diarrhoea, GE, GBS, ReA, IBS, Death | 0.029 |
|  | de Noordhout *et al.*, 2017 (51) | No | No | Country specific; and GBD 2010 | Incidence | Literature | European DW | GE, GBS, ReA, death | 1.609, 1.605 (2012 and 2020 predictions) |
|  | Mangen *et al.*, 2015 (42) | Yes | No | West Level 25 and 26 | Incidence | Expert opinion | Dutch DW | GE, GBS, ReA, IBD, IBS, Death | - |
|  | Lai *et al.*, 2020 (52) | No | No | Country-specific | Incidence | Literature and expert opinion | GBD 2016 and WHO/FERG | Diarrhoea, GE, Death | - |
|  | van Lier *et al.*, 2016 (43) | No | No | West Level 26 | Incidence | Literature and expert opinion | DW compiled from: GBD, Person-Trade-Off, and novel elicitation methods | GE, GBS, ReA, IBD, IBS, Death | 0.492, 0.515, 0.456, 0.399, 0.387 (2007-2011 respectively) |
|  | Havelaar *et al.*, 2012 (44) | Yes | No | West Level 25 and 26 | Incidence | Literature | Dutch DW | GE, GBS, ReA, IBD, IBS, Death | - |
|  | Haagsma *et al.*, 2010 (45) | No | No | N/A | Incidence | Literature | IBS DW | IBS | 0.038, 0.020 (including and excluding PI-IBS respectively) |
|  | Gibney *et al.*, 2014 (53) | No | Yes | Country-specific | Incidence | Literature | DW compiled from: GBD 2010 and Dutch DW | GE, GBS, ReA, IBS, Death | - |
|  | Martins *et al.*, 2017 (54) | No | No | Country-specific | Incidence | Literature and data | GBD 2010 | GE (diarrhoea), GBS, ReA, IBD, IBS, Death | 0.057, 0.061 (best-case scenario for 2008 and 2013) |
|  | Sapp *et al.*, 2023 (59) | No | No | WHO 2050 | Incidence | Literature and data | GBD 2010 | Diarrhoea, GBS, Death | - |
|  | Kumagai *et al.*, 2015 (55) | No | No | GBD 2010 | Incidence | Literature | European DW | GE, GBS, ReA, IBD, Death | 0.051 |
|  | Pires *et al.*, 2022 (50) | No | No | Country-specific | Incidence | Data | GBD 2013 | Diarrhoea, GE, GBS, ReA, IBS, Death | 0.028 |
|  | Havelaar *et al.*, 2000 (46) | No | No | GBD 1996 | Incidence | Literature | GBD 1996 | GE, GBS, ReA, Death | - |
|  | Toljander *et al.*, 2012 (56) | No | No | Country-specific | Incidence | Literature and data | Dutch DW | GE, GBS, ReA, Death | 0.004, 0.001, (deterministic scenarios) |
|  | Gkogka *et al.*, 2011 (57) | No | No | Country-specific and West | Incidence | Literature | DW compiled from: IBS DW; GBD | GE, GBS, ReA, IBD, IBS, Death | - |
|  |  |  |  | Level 26 |  |  |  |  |  |
|  | Mangen *et al.*, 2005 (47) | Yes | No | Country-specific | Incidence | Literature | Dutch DW | GE, GBS, ReA, IBD, Death | - |
|  | Lackner *et al.*, 2019 (58) | No | No | Country-specific | Incidence | Literature | European DW | GE, GBS, ReA, IBD, IBS, Death | 0.001, 0.180, 0.496, 7.747, 8.817 (GE, ReA, IBS, GBS and IBD respectively) |
|  | Mangen *et al.*, 2013 (48) | No | No | West Level 25 and 26 | Incidence | Literature | GBD DW | GE, GBS, ReA, IBS, Death | - |
| *GBD data for campylobacteriosis does not exist as a standalone disease as it is considered as an aetiology of overall infectious diseases, hence, information on morbidity and mortality is combined with several other diseases.<br>GE = gastroenteritis; IBS = irritable bowel syndrome; IBD = inflammatory bowel disease; ReA = reactive arthritis; GBS = Guillain-Barré syndrome |  |  |  |  |  |  |  |  |  |

#### Approaches for Estimating YLD: Incidence- vs. Prevalence-Based Approach

Most studies (n=22) utilised the incidence-based approach to compute YLDs associated with campylobacteriosis (15,26,31,40,42–59), while the GBD study by Vos *et al.,* (41) employed the prevalence-based approach. Some studies addressed comorbidity to enhance burden estimates. For instance, Mangen *et al.,* (47) adjusted for comorbidity by applying a correction factor of 1.86 across all ages. This factor was derived from a Danish study (60) that estimated the relative risk of mortality for patients with laboratory-confirmed campylobacteriosis within the first year after symptom onset. The estimation utilized an index based on relative mortality rates from different diagnostic groups. This adjustment revealed a relative risk of 1.86 (90% CI 1.56–2.20), accounting for other health conditions. Similarly, Kirk *et al.*, (15) included a comorbidity adjustment in their global assessment of 22 diseases, however, their study did not specifically address the burden associated with campylobacteriosis.

#### Pathogen- vs. Outcome-Based Approach

The majority of studies (91%) utilised the pathogen-based approach, many of which also employed the incidence-based approach. Notably, two recent publications studies by Lai *et al.,* (52) and van Lier *et al.,* (43) reported adopting the WHO/FERG and BCoDE protocols, respectively. It is important to note that the WHO/FERG (37) study was not directly included in the data abstraction process because it did not provide specific BoD estimates. Instead, it focused on demonstrating how the methodology can be used to convert epidemiological data into BoD estimates (37). In contrast, two studies followed an outcome-based approach; one of these was the GBD study (41). Diverging from the GBD methodology, Haagsma *et al.,* (45) applied both an incidence and outcome-based approach, specifically examining the disease burden of post-infectious IBS following campylobacteriosis by calculating the number of IBS cases attributable to *Campylobacter* infection.

### Health States

Fig 3 presents the perceived outcome tree for campylobacteriosis, adapted from Mangen *et al.*, (48) and Lackner *et al.,* (58). However, there was notable variation in the health states associated with campylobacteriosis, leading to the application of seven different disease models across studies (S4 Campylobacteriosis Disease Models). Among studies calculating DALYs for campylobacteriosis, seven studies reported various health states including gastroenteritis, IBS, IBD, ReA, GBS, and mortality (disease model 1) (26,42–44,54,57,58). Nonetheless, Martins *et al.,* (54) considered cases of gastroenteritis in conjunction with diarrhoea. Conversely, some studies (15,49,50,52,59) treated diarrhoea as a distinct health state from gastroenteritis, including individual-level national (n=4) and global studies (n=1). While most studies reported all health states and their sequelae, some omitted IBD (31,40,48–50,53) or IBS (47,55), or both (46,51,56). Haagsma *et al.,* (45) exclusively considered post-infectious IBS following *Campylobacter* infection and did not compute YLLs since IBS was not associated with increased mortality (disease model 7).

**Fig 3.**
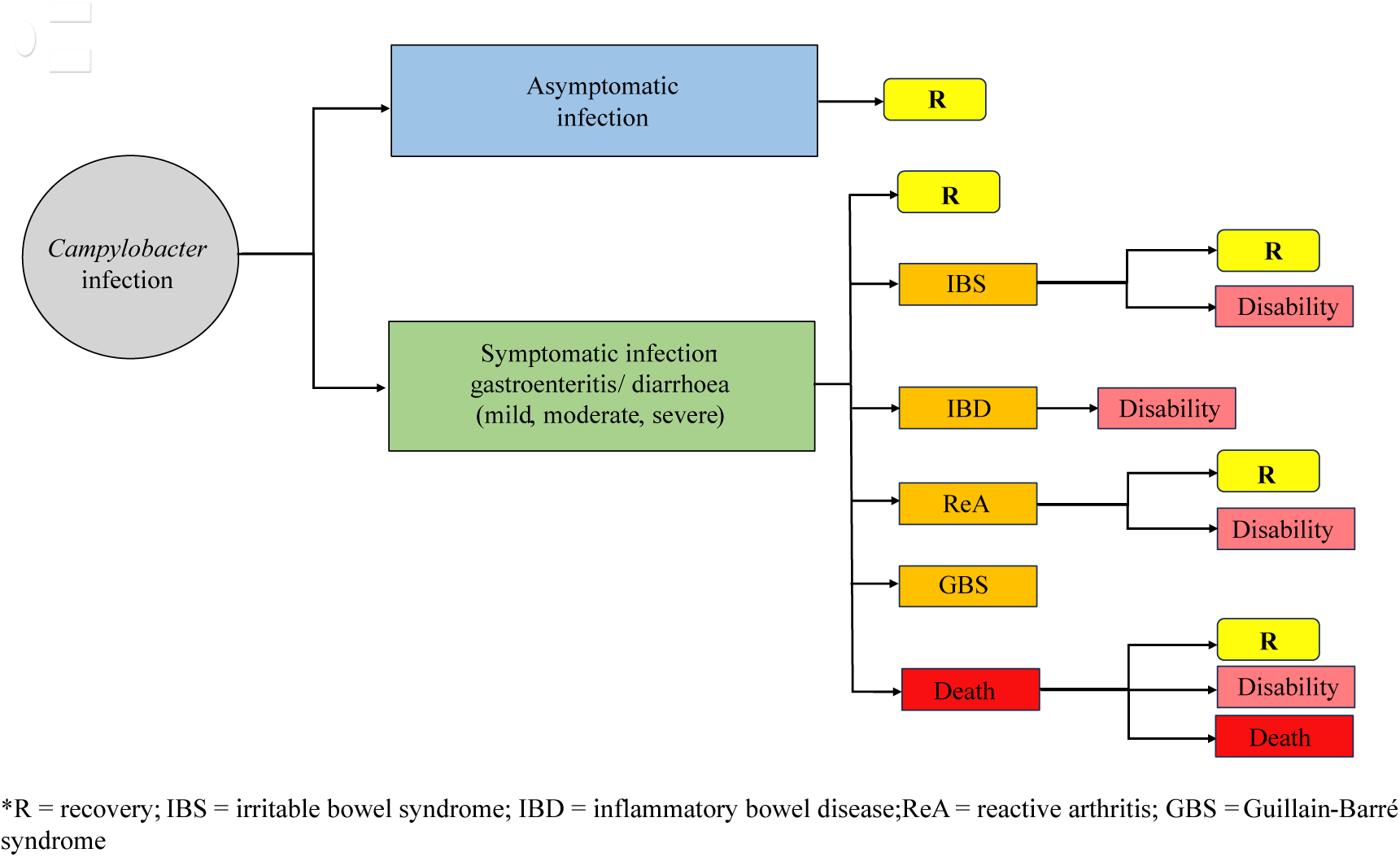
Illustration of the perceived outcome tree for *Campylobacter* infection in humans (adapted from Mangen *et al.*, (48) and Lackner *et al.,* (58)).

### Choice of Disability Weights

Based on the 23 studies, the GBD DW was the most commonly used (39%) (15,40,41,46,48–50,54,59). Among these, the DWs largely stemmed from the GBD 2010 study (15,54,59). Notably, Martins *et al.,* (54) and Kirk *et al.,*(15) computed DWs based on the GBD 2010 framework, albeit with variances in the classification of health states. For instance, Kirk *et al.,* (15) categorized certain clinical outcomes for campylobacteriosis into acute *Campylobacter* diarrhoea (mild), acute *Campylobacter* diarrhoea (moderate), and acute *Campylobacter* diarrhoea (severe), assigning DWs of 0.061, 0.202, and 0.281, respectively. Similarly, Martins *et al.,* (54) adopted these DWs as proposed by GBD 2010 but considered gastroenteritis alongside diarrhoea, analysing them as mild, moderate, or severe, following the same pattern. In contrast, the European DW set, recommended and implemented by the BCoDE study, conducted by Cassini *et al.,* (31) were also included in three other studies. These national-level studies conducted in Japan (55), Belgium (51), and Germany (58), published in 2015, 2017, and 2019 respectively, after the elicitation of the European DWs in 2015. Several studies opted for alternative DWs, primarily the Dutch DWs (26,42,44,47,56). The majority of studies (n=3) employing the Dutch DWs were conducted at a national level within the Netherlands (42,44,47). Conversely, two national studies, in Sweden (56) and the United States (26) also used the Dutch DWs. The remaining studies either used specific DWs for sequelae (45), or amalgamated existing DW sets (43,52,53,57).

### Duration

Data concerning the duration of disease were obtained from various sources, including existing literature, data analysis, and expert opinion (Table 1). However, one study by Scallan *et al.,* (26) lacked clarity regarding this process. Additionally, as campylobacteriosis is not considered a standalone cause of morbidity and mortality in the GBD study (41), data extraction was not feasible. Among the studies reviewed, the majority (n=13) relied solely on existing literature to define health states and duration data (15,31,40,44–48,51,53,55,57,58). However, significant variations emerged regarding the duration of sequelae associated with campylobacteriosis (44–46,48,53). For example, Gibney *et al.,* (53) reported durations of 0.6 years, 5 years, and lifelong for ReA, IBS, and GBS, respectively. Mangen *et al.,* (48) similarly concluded that IBS could persist for 5 years as well as severe cases of GBS lasting for an individual’s remaining life expectancy. They did, however, state that GE could only persevere for 0.017 (years) (48). In contrast, Havelaar *et al.,* (46) suggested a shorter duration for GE (5.08 days) and ReA (6 weeks), but surprisingly, did not consider GBS as a lifelong disability, estimating its duration at 1 year. Conversely, another study by Havelaar *et al.,* (44) assumed IBD to be lifelong, while Haagsma *et al.,* (45) indicated that symptoms of post-infectious IBS could last for 5 years.

Alternatively, two independent studies in Denmark estimated campylobacteriosis duration through data analysis (49,50). Pires *et al.,* (49) based their findings on life expectancy data provided by Denmark Statistics. They determined that symptoms of diarrhoea typically lasted for 3 days, while ReA persisted for 222 days. In contrast, both IBS and GBS were identified as severe, long-lasting conditions, with IBS potentially lasting for 1825 days and GBS extending for the duration of an individual’s remaining life expectancy (49). Mangen *et al.,* (42) based their duration data on expert elicitation. The remaining studies (n=5) employed a combination of methods, drawing from published literature, data analysis, and expert opinion (43,52,54,56,59). Toljander *et al.,* (56), for instance, synthesized duration data from literature, national notification systems, and hospitalization records, establishing that reported GE typically lasted for 8.37 days, ReA for 0.71 years, short-term (≤ 1 year) GBS for 0.685 years, and long-term (> 1 year) GBS for 15.2 years.

### Epidemiological Data and Data Adjustments

Based on the included studies, several epidemiological data sources were utilized to calculate DALYs. The majority of studies (n=9) drew information from published literature (15,31,45,47,48,52,53,56,59). Moreover, three studies combined literature findings with national surveillance data (15,47,56), one study relied on sentinel surveillance (48), and another cited the WHO/FERG database (59). Notably, two studies incorporated data from multiple sources, with Gibney *et al.,* (53) integrating national estimates from administrative data and unpublished data from OzFoodNet, while Lai *et al.,* (52) gathered information from various Taiwanese authorities, national databases, and nationwide surveys. In contrast, five studies predominantly obtained incidence data from laboratory-confirmed cases (40,43,46,50,51). Each of these studies enhanced their findings by incorporating supplementary data from various sources, including sentinel surveillance, statutory notification data, and reports from outbreaks, population-based surveys, and general practitioner surveillance (40,43,46,50,51). Notably, the studies from Scallan *et al.,* (26), Pires *et al.,* (49), Mangen *et al.,* (42), Havelaar *et al.,* (44), and Gkogka *et al.,* (57), sourced data from multiple inputs, including published literature, laboratory cases, active surveillance, and national medical registers. Furthermore, three studies explicitly utilized alternative data sources, such as those from the Federal Office of Public Health or local government statistics, registration records, and national patient surveys (54,55,58).

To mitigate underestimation of incidence, fourteen studies adopted data adjustment techniques. Five studies carried out data adjustments through a data-driven approach (26,46,52,55,57). Lai *et al.,* (52) adjusted for underestimation by selecting the highest estimate as the most accurate reflection of the actual incidence of diagnoses, while Gkogka *et al.,* (57) employed a PERT distribution derived from previously published literature as multipliers to account for food attribution. Alternatively, others relied on expert opinions to manage underestimation (40,43,48,49,51,53,54). Some of these studies (40,43,48,51) based their adjustments on guidelines outlined by Havelaar *et al.,* (61). The remaining studies also adopted an expert-driven approach, albeit with differing methodologies (49,53,54). Gibney *et al.,* (53) utilized regional data to estimate the number of notifiable cases of *Campylobacter* in Australia, complemented by case fatality ratios from a study on FBDs in the United States (26). Similarly, Pires *et al.,* (49) tailored the surveillance pyramid outlined by Haagsma *et al.,* (62) to adjust for underreporting and underdiagnosis of diseases. Only one study (50) combined both approaches, incorporating expert opinions and non-pathogen-specific and pathogen-specific parameters to adjust for underreporting and underdiagnosis. Additionally, several studies (n=13) employed Monte Carlo simulation to model uncertainty (15,31,42–45,48,51,53,55,56,58,59).

## Discussion

This systematic review aimed to comprehensively assess the existing studies on the burden of campylobacteriosis in humans, focusing on methodological designs and assumptions used in calculating DALYs across studies published between 1990 and July 2023. The inclusion criteria prioritized studies quantifying DALYs attributable to campylobacteriosis, regardless of their primary research focus, ensuring a comprehensive analysis of the disease burden. Methodologically, the reviewed studies employed diverse approaches including both incidence-based and prevalence-based approaches, which influenced the calculation and interpretation of DALYs.

Among the 23 studies that met the inclusion criteria, many emphasized single-country analyses, with the Netherlands significantly represented (n=7). Although zoonotic and FBDs like *Campylobacter* hold global significance, regional variations in data availability and research efforts have primarily focused on higher-income countries, resulting in a notable lack of understanding the burden on marginalized populations and LMICs. The scarcity of available data from LMICs is evident, with only one estimate from Rwanda, noted by Sapp *et al.*, (59). Despite being recently published, the study’s data pertains to the reference year 2010, underscoring the ongoing challenge in data acquisition from these regions.

Our findings revealed varying methodological approaches across studies, particularly concerning life expectancy tables, which greatly influences the YLL and international comparability. The decision to use local or universal life expectancy tables is pivotal, with national life tables commonly utilised by many national BoD studies such as those from the United States of America (26), Taiwan (52), Australia (53), Sweden (54,56), Denmark (50), the Netherlands (47), and Germany (58). However, the lack of established standards for these tables hinders cross-country comparability (33). Conversely, aspirational model life tables, like those used in the GBD study, are often referred to in the literature as the optimal standard (30,33). However, the use of a universal life expectancy, which is intended to ensure that the death of an individual at a given age is weighted equally across different countries and contexts, faces criticism due to potential confounding variables across different socioeconomic regions (30). Moreover, the GBD methodology excludes stillbirths from the DALY model (30,63), an issue relevant for campylobacteriosis, as certain strains like *C. jejuni*, have been associated with septic abortion in pregnant women following aggressive bowel infections (2). This connection adds complexity to determining the *“age at which DALYs should be counted”*, as even minor adjustments in DALY calculations can significantly impact disease burden assessments (63).

The integration of age weighting and time discounting for DALY estimates has long been controversial, often referred to as *“the devils in the DALY”* (33,63,64). First used in the GBD 1990 study, these parameters have been criticized for underestimating the BoD by reducing YLLs due to premature death (63). Since 2010, the GBD has omitted social weighting, with many BoD studies following suit (65). Indeed, 94% of studies that excluded social weighting were published after 2010, reflecting this methodological shift. Despite this trend, national BoD studies often continue to use these parameters, suggesting ongoing methodological debate. This review identified a small number of studies that included time discounting or social weighting, indicating a lack of consensus regarding their use in BoD assessments (33).

Methodological diversity was also evident in the choice between prevalence- and incidence-based approaches for estimating disease burden (66). While the prevalence-based approach has gained prominence in recent years, the incidence-based approach remains preferred, particularly in studies concerning FBDs. The debate between these approaches centres on how morbidity is allocated within a given year (63). The incidence-based approach assigns all health states, including future ones, to the current event, while the prevalence method reflects the current BoD from past events (25,63). Despite the GBD study’s transition, this review highlights the widespread use of the incidence-based approach in BoD studies spanning both single- and multi-country analyses. Our findings emphasize its suitability for foodborne and infectious diseases, attributed to its heightened sensitivity to epidemiological data and its effectiveness in capturing YLD, particularly for short duration diseases like campylobacteriosis (29,37,66).

Most of the BoD assessments that utilized the incidence-based approach also predominantly used a pathogen-based approach. Among these, two studies explicitly mention adhering to the WHO/FERG and BCoDE frameworks. However, the WHO/FERG (37) study was not included in the data abstraction process of this review because it did not provide specific BoD estimates, focusing instead on developing methodologies. In contrast, the GBD study employs a prevalence-based approach alongside an outcome disease model for presenting fatal and non-fatal events (41). While various methodologies exist for quantifying DALYs, the WHO/FERG methodological framework suggests aligning the methodological approach with the nature of the disease under investigation, be it hazard, outcome, or risk factor (37). Given campylobacteriosis is a FBD associated with multiple hazards, the pathogen-based approach is often deemed more suitable (37).

Establishing DWs lacks a universal standard, complicating the assessment of their validity (38,67). This variation in DW selection adds complexity to comparisons between studies, with both GBD DWs and alternative DWs, like Dutch and European DWs facing criticism. Although GBD DWs are widely used, Havelaar *et al.,* (46) applied the older GBD 1996 DWs. These DWs, derived from expert evaluations of various health states, have been criticized for inconsistencies among experts and for not adequately representing global health states (30,33). Revisions to the GBD methodology have sought to address these concerns (33,65). Charalampous *et al.,* (38) noted that panels involving the general public must ensure that health descriptions are comprehensible to laypersons, as their lack of disease expertise can compromise the validity of health state valuation data. The use of alternative DWs, such as Dutch and European DWs, also complicates the interpretation of BoD studies. While they may offer insights into morbidity severity within specific cultural contexts, they hinder the comparability and validity of results across different studies (68).

Considerable heterogeneity exists in defining health states associated with campylobacteriosis, particularly regarding the inclusion of IBD or IBS as sequelae. While earlier studies frequently included IBD as a sequelae of campylobacteriosis, more recent publications have tended to (31,40,46,48–51,53,56). Pires *et al.,* (49) also noted this trend, suggesting that the perceived association between *Campylobacter* infection and IBD might be overestimated, with detection bias from recurrent stool testing likely contributing to its occurrence, rather than true causality (49,69). The varied symptoms, complications, and long-term effects of *Campylobacter* infection further complicate comparisons of health states, especially given discrepancies in the literature regarding its pathogenicity (26).

Estimating the duration of campylobacteriosis’ burden entails significant methodological variability, drawing from diverse data sources. This variability is evident in the DALYs per case reported in various studies, highlighting the impact of methodological choices and assumptions on the overall DALYs per case result. For instance, Toljander *et al.,* (56) reported the lowest DALYs per case result by excluding social weighting and employing Dutch DWs. In contrast, Mangen *et al.,* (40) also reported relatively low DALYs per case by utilizing GBD DWs and employing various time discounting rates. These disparities underscore the lack of methodological consistency and substantial variations in results, cautioning against straightforward interpretation of BoD estimates, especially when comparing them internationally.

Most of the BoD studies examined were conducted independently, primarily in Western Europe, suggesting a correlation between surveillance intensity and economic status (36). High-income nations benefit from abundant data availability and timely reporting, enhancing the quality of surveillance systems. In contrast, LMICs face challenges in achieving similar surveillance standards, leading to delays in epidemiological data publication and hindering accurate depiction of campylobacteriosis incidence, mortality, and overall burden (2,36,70). Most studies sourced epidemiological data from previously published literature. Those that primarily depended on laboratory-confirmed cases for incidence evaluation also included additional data sources. Relying solely on laboratory-confirmed data may underestimate results due to the potential exclusion of mild or asymptomatic cases that go unreported, compounded by variations in healthcare-seeking behaviour, reporting systems, and diagnostic practices across populations and settings (1,28). Consequently, many studies integrated laboratory-confirmed cases with other data sources, resulting in wider uncertainty ranges in BoD estimates and influencing the interpretation of findings. Most studies employed Monte Carlo simulations as a method to assess uncertainty. However, determining the most appropriate model for BoD studies remains variable, reflecting the diverse nature of diseases and their impacts across different populations.

As zoonotic and FBDs pose ongoing social and economic burdens (3,71), improving the accuracy and policy relevance of DALY estimates is essential, particularly for diseases like campylobacteriosis, a prevalent but often *“neglected foodborne zoonotic pathogen”* (1,18). However, methodological inconsistencies in disease burden estimation hinder public health efforts and One Health objectives (72), which seek interdisciplinary synergy among human, animal, and environmental health. This underscores the need for refined methodologies to ensure accurate and applicable BoD estimates for effective public health research and policymaking.

### Limitations

This review has several limitations. Firstly, a risk of bias assessment was not performed for the included studies, as it is not deemed suitable for evaluating BoD studies. Additionally, despite efforts to retrieve relevant literature from various databases and platforms, there is a possibility of overlooking certain grey literature sources. Notably, only a single study was identified from a LMIC, where the burden of campylobacteriosis may be higher but remains unaccounted for in this review.

### Conclusions and Future Prospects

This systematic review aimed to examine the existing literature on *Campylobacter* BoD studies, focusing on the methodological approaches used to quantify DALYs. This systematic synthesis of evidence suggests that many of these studies predominantly utilized an incidence-based approach, frequently accompanied by a pathogen-based method, aimed at comprehending the progression of campylobacteriosis and its associated outcomes. Nevertheless, notable disparities were evident concerning several parameters, encompassing the selection of DWs, disease duration, and health state categorization.

Despite the growing importance of BoD studies for population health and resource allocation, their complex methodologies pose challenges. This review underscores the need for methodological rigor and harmonization to advance future BoD studies employing the DALY approach. Improving comparability and validity rests upon addressing data gaps and establishing consensus on which sequelae to include in DALY calculations, considering the observed variability in health state values associated with campylobacteriosis. Failure to address these methodological constraints could compromise BoD estimates, hindering cross-country comparisons and public health efforts. Collaborative efforts are essential to ensure the robustness and relevance of BoD studies in informing policies and interventions.

## Data Availability

All relevant data are within the manuscript and its Supporting Information files.

## Author Contributions

**Conceptualization:** Megan Tumulty, Zubair Kabir, Carlotta Di Bari, Brecht Devleesschauwer

**Data Curation:** Megan Tumulty

**Formal Analysis:** Megan Tumulty, Zubair Kabir, Carlotta Di Bari

**Funding Acquisition:** Not applicable

**Investigation:** Megan Tumulty, Zubair Kabir, Carlotta Di Bari

**Methodology:** Megan Tumulty, Zubair Kabir, Carlotta Di Bari, Sara Monteiro Pires

**Project Administration:** Megan Tumulty, Zubair Kabir, Carlotta Di Bari

**Resources:** Not applicable

**Software:** Not applicable

**Supervision:** Zubair Kabir, Carlotta Di Bari

**Validation:** Megan Tumulty, Zubair Kabir, Carlotta Di Bari, Brecht Devleesschauwer, Sara Monteiro Pires

**Visualization:** Megan Tumulty

**Writing – Original Draft Preparation:** Megan Tumulty

**Writing – Review & Editing:** Megan Tumulty, Zubair Kabir, Carlotta Di Bari, Brecht Devleesschauwer, Sara Monteiro Pires

## Supporting Information

**S1 PRISMA Checklist**

**S2 Search Strategy**

**S3 Data Abstraction Form**

**S4 Campylobacteriosis Disease Models**

**S5 Disability-Adjusted Life Years Calculations**

